# SARS-CoV-2 seroprevalence trends in healthy blood donors during the COVID-19 Milan outbreak

**DOI:** 10.1101/2020.05.11.20098442

**Authors:** Luca Valenti, Annalisa Bergna, Serena Pelusi, Federica Facciotti, Alessia Lai, Maciej Tarkowski, Alessandra Berzuini, Flavio Caprioli, Luigi Santoro, Guido Baselli, Carla della Ventura, Elisa Erba, Silvano Bosari, Massimo Galli, Gianguglielmo Zehender, Daniele Prati

## Abstract

**Background&Aims:** The Milan metropolitan area in Northern Italy was among the most severely hit by the SARS-CoV-2 outbreak. The aim of this study was to examine the seroprevalence trends of SARS-CoV-2 in healthy asymptomatic adults, the risk factors, and laboratory correlates.

**Methods:** We conducted a cross-sectional study in a random sample of blood donors since the start of the outbreak (February 24^th^ to April 8^th^ 2020, n=789). Presence of IgM/IgG antibodies against the SARS-CoV-2-Nucleocapsid protein was assessed by a lateral flow immunoassay.

**Results:** The test had a 100/98.3 sensitivity/specificity, and for IgG+ was validated in a subset by an independent ELISA against the Spike protein (N=34, *P*<0.001). At the outbreak start, the overall adjusted seroprevalence of SARS-CoV-2 was 2.7%, 95% c.i. 0.3-6% (*P*<0.0001 vs. 120 historical controls). During the study period characterized by a gradual implementation of social distancing measures, there was a progressive increase in adjusted seroprevalence to 5.2%, 95% c.i. 2.4-9.0, due to a rise in IgG+ tests to 5%, 95%CI 2.8-8.2 (*P*=0.004 for trend, adjusted weekly increase 2.7±1.3%), but not of IgM+ (*P*=NS). At multivariate logistic regression analysis, seroconversion to IgG+ was more frequent in younger (*P*=0.043), while recent infections (IgM+) in older individuals (*P*=0.002). IgM+ was independently associated with higher triglycerides, eosinophils, and lymphocytes (*P*<0.05).

**Conclusions:** SARS-CoV-2 infection was already circulating in Milan at the outbreak start. Social distancing may have been more effective in younger individuals, and by the end of April 2.4-9.0% of healthy adults had evidence of seroconversion. Asymptomatic infection may affect lipid profile and blood count.

## INTRODUCTION

The Milan metropolitan area in Northern Italy was the first, among those in Western countries, severely hit by the spread of SARS-CoV-2, which is the virus that causes COVID-19, with a 26% mortality rate in critically ill patients [1].

A fraction of individuals infected by SARS-CoV-2 are asymptomatic or mildly symptomatic and they represent a major source of viral spread [2, 3, 4]. It has been estimated that 9.8% of Italian population has already been infected by SARS-CoV-9 [5]. However, epidemiological trends in individuals with mild COVID-19 remain unknown. SARS-CoV-2 replication is followed by IgM/IgG seroconversion, which can improve COVID-19 diagnosis and the evaluation of disease circulation [6, 7]. On the other hand, it is not known whether routine laboratory tests may help identifying asymptomatic carriers.

The Transfusion center at the Fondazione IRCCS Ca’ Granda Policlinic Hospital is the main blood center in Milan, collecting almost 40,000 blood donations per year. During the last 25 years, we provided evidence that blood donor cohorts represent a special vantage point to study subclinical conditions, and to describe the prevalence, incidence and natural course of infectious diseases [8, 9]. Studies in blood donors might help to assess the dynamics of viral circulation, and for modelling the evolution of theCOVID-19 outbreak [10].

The main aim of this study was therefore to examine the trend in the prevalence of SARS-CoV-2 antibody reactivity among healthy and asymptomatic individuals during the outbreak, the risk factors, and laboratory features associated with recent infection.

## METHODS

### Study cohorts

We considered 789 individuals donating blood between February 24^th^ (the first week of the Italian outbreak) and April 8^th^ 2020, whose plasma samples were stored for hemovigilance studies.

The main study cohort was composed of blood donors, who were apparently healthy subjects, aged 18-70 years. Exclusion criteria were any active infection or other active medical conditions, recent surgical procedures, trips in areas with endemic infective diseases, reported risk factors for parenterally acquired infections, chronic degenerative conditions except stable arterial hypertension, type 2 diabetes or dyslipidemia under control with lifestyle or pharmacological therapy, diagnosis of cancer or high risk of cardiovascular events. All donors underwent clinical and medical history evaluation and biochemical testing. To qualify for blood donation, candidates should had been free of recent symptoms possibly related to COVID-19, nor had close contact with confirmed cases. Since March 26^th^, they should had been symptoms free during the preceding 14 days, nor had unprotected contacts with suspected cases [11].

Among the 3,586 individuals who donated blood during the study period, we randomly selected 20 per each day to assess SARS-CoV-2 seroprevalence, whose clinical features were representative of the overall population (not shown). Clinical features of 789 included individuals are shown in Table 1.

**Table 1.** Clinical features of 789 individuals, who donated blood between February 24^th^ and April 8^th^, stratified by the presence of anti-SARS-CoV-2 antibodies.

|  | SARS-CoV-2 antibodies |  |  |  | P-value* |
| --- | --- | --- | --- | --- | --- |
|  | Present |  | Absent |  |  |
| N= | 40 | (5.1) | 749 | (94.9) |  |
| Age, years | 42.6±13.4 |  | 40.7±13.2 |  | 0.37 |
| Sex, F | 10 | (25.0) | 266 | (35.6) | 0.16 |
| Residency, Milan | 24 | (60) | 497 | (66.4) | 0.91 |
| Health-care profession, yes | 2 | (5) | 35 | (5.1) | 0.75 |
| BMI, Kg/m<sup>2</sup> | 25.5±3.3 |  | 24.3±3.7 |  | 0.040 |
| Arterial hypertension, yes | 2 | (3.2) | 60 | (8.1) | 0.76 |
| Smoking, active | 7 | (17.5) | 197 | (27.3) | 0.27 |
| ABO blood group:<br>o/A/A1/AB/B | 16/16/0/2/6 | (40/40/0/5/15) | 341/287/3/27/90 | (45.6/38.4/0.4/3.6/12.0) | 0.92 |
| Week of outbreak, n | 4.5 | [2-6] | 4.0 | [2-5] | 0.10 |
| Lymphocytes, 10<sup>3</sup>/mm<sup>3</sup> | 1.94±0.54 |  | 1.91±0.52 |  | 0.70 |
| ALT, IU/l | 25.3±11.9 |  | 25.6±16.0 |  | 0.81 |
| GGT, IU/l | 24.3±20.0 |  | 18.2±13.1 |  | 0.090 |

Data are shown as mean±SD, median [IQR], N (%) values. ALT: alanine aminotransferases, GGT: gamma-glutamyl transferases. \* At logistic regression analysis.

To gain further insight into the epidemiological trends before the outbreak, we also examined anonymized samples of 184 individuals, who presented for blood donation between December 2019 and March 2020 and were included in a screening program for metabolic disorders (Bible study, mean age 54.7±6.4, 89.6% of male sex, body mass index (BMI) 28.8±3.4 Kg/m^2^).

The study protocol complies with Good Clinical Practice (GCP) rules, Declaration of Helsinki, European clinical practice, international guidelines and national law regulation in Italy, and was approved by the Ethical Committee of the Fondazione IRCCS Ca’ Granda (“COVID-19 Donors Study”, CoDS), n.334-2020 on 03/04/2020. Each blood donor signed written informed consents allowing for testing for communicable diseases, storage of anonymized data and biological materials for diagnostic and research purposes, and use of their de-identified data for clinical research.The blood donors’ organization supporting our center was involved in the study planning and design (https://www.donatoriamici.it). The study participants were involved in the research at the time when the study was presented and informed consent signed. Implementation of widespread testing of donors during the next study phase will be implemented in concern with the blood donors’ organization.

### Evaluation of anti-SARS-CoV-antibodies

The presence of IgM/IgG against SARS-CoV-2 were determined on plasma samples (20 μl) by a lateral flow immunoassay against the Nucleocapsid protein (COVID-19 IgG/IgM Rapid Test, Prima Lab, Balerna, CH). The study test was chosen because at the time of protocol approval had already obtained CE certification and in pre-clinical evaluation provided reasonable accuracy when compared to RT-PCR results, and it was performed at the Laboratory of Infectious Diseases at “Luigi Sacco” Hospital. The antibody is directed against the nucleocapsid antigen of SARS-CoV-2. The reported accuracy for the lateral flow immunoassay, were for IgG: specificity 98.0%, sensitivity 100%, accuracy 98.6%; for IgM: specificity 96.0%, sensitivity 85.0%; accuracy 92.9%. In addition, it was easy to perform and potentially applicable to rapid pre-donation screening. The test was read by two independent expert biologists, who had to reach an agreement, and considered positive when strong or weak immunoreactivity for IgG or IgM was detected, while very weak or dubious positive results were considered not specific. The serologic assay was validated by testing the plasma of 22 COVID-19 patients (confirmed by qRT-PCR) 10 days after they were hospitalized in Milan between February and March 2020 (positive controls). A total of 36 serum samples have been collected at different times from the onset of symptoms. As negative controls we examined 120 patients who were assessed for blood transfusion during the year 2009 at the Policlinic hospital in Milan.

In a subset of cases (n=34, 5 IgG positive (IgG+), 5 IgG/IgM+, 4 IgM+, 21 negative) the lateral flow immunoassay IgG test was validated against a home-made ELISA test evaluating antigens in the SARS-CoV-2 spike protein [11]. The diagnostic threshold for positive results of ELISA IgG (OD=0.386) was selected on the basis of a receiver operating characteristic (ROC) curve analysis performed on historical negative controls and in an independent set of swab-confirmed COVID-19 cases (100% sensitivity and 97% specificity).

### Statistical analysis

We assumed that the outbreak of SARS-CoV-2 infection started at the beginning of February 2020 in the Milan area, so that we could expect a low rate (95% confidence interval (c.i.) 0-2%) of IgM+ at the beginning of the study (end of February 2020). We report here the results of analysis of the first CoDS study period, which was pre-planned to gain a timely insight into the dynamic of viral spread to inform healthcare decisions.

For descriptive statistics, continuous traits were summarized as mean±SD, while highly skewed variables were summarized as medians and interquartile range. Categorical variables were shown as percentages. The seroprevalence was reported as rate and 95% c.i.. We used these data to estimate the population prevalence of SARS-CoV-2 antibodies. The proportion of positive tests (either IgM or IgG) in the analysis was then adjusted for the diagnostic accuracy of the test. We also provided a further estimation of prevalence of diseases adjusted for the accuracy of the test by a Bayesian approach [12]. Estimates of sensitivity and specificity of the test were derived from local cases (n=22) and controls (n=120); we used the 95% confidence intervals for Bayesian estimation of SARS-CoV-2 seroprevalence. Analyses were performed by fitting data to logistic regression models were fit to examine binary traits (presence of IgM and/or IgG antibodies; see also the supplementary). Analyses were adjusted for main known confounders, as specified in the results section. *P*-values <0.05 (two tailed) were considered statistically significant.

Results were reported according to the STROBE guidelines. Statistical analysis was carried out using the JMP Pro 14.0 Statistical Analysis Software (SAS Institute, Cary, NC), and R statistical analysis software version 3.5.2 (http://www.R-project.org/).

## RESULTS

### Validation of the diagnostic test

The lateral flow immunoassay showed a 100% sensitivity for IgG (95% c.i. 84-100%), 68% sensitivity for IgM (95% c.i. 45-86%) for detecting SARS-CoV-2 infection in the 22 COVID-19 patients 10 days after they were hospitalized (Figure S1). In addition, the test detected a clear positivity for IgG in one mildly symptomatic individual, who was tested two weeks after viral RNA on nasal swab became negative. In 120 patients who were assessed for blood transfusion during the year 2009, the specificity was 98.3%, 95% c.i. 94.1-99.5% (99.2% for both IgM+ and IgG+). The two false positive individuals were in their sixties and had a recent history of cancer, which was active urological cancer in a male with IgG+, while the IgM+ woman had a diagnosis of rheumatoid arthritis with positive rheumatoid factor.

The lateral flow immunoassay had a good agreement with ELISA test to detect IgG (k=0.59±0.16, *P*<0.001). Interestingly, 3 out of 5 discordant cases between lateral flow and ELISA test occurred in IgM+ samples, suggesting that IgM+. In IgM negative samples, the agreement of lateral flow immunoassay and ELISA IgG increased to k=0.71±0.19, *P*<0.001).

### Seroprevalence trends during the outbreak

The trend in the overall seroprevalence, IgM, IgG and the combined profile is reported in Figure 1. During the first two weeks, the baseline prevalence was 4.6% (2.3 to 7.9%; *P*<0.0001 vs. historical controls; 3.7%, and 2.0% for IgM+/IgG+, respectively). After adjustment for imperfect accuracy of the test, the estimated prevalence was 2.7%, 95%CI 0.3-6%. During the study (Figure 1A), there was a trend for an increase in the overall seroprevalence to 7.1% (*P*=0.036), due to an increase in IgG+ (*P*=0.019). We observed a tendency for reduction in the prevalence of IgM+ to 1.7%, which was not significant.

**Figure 1.**
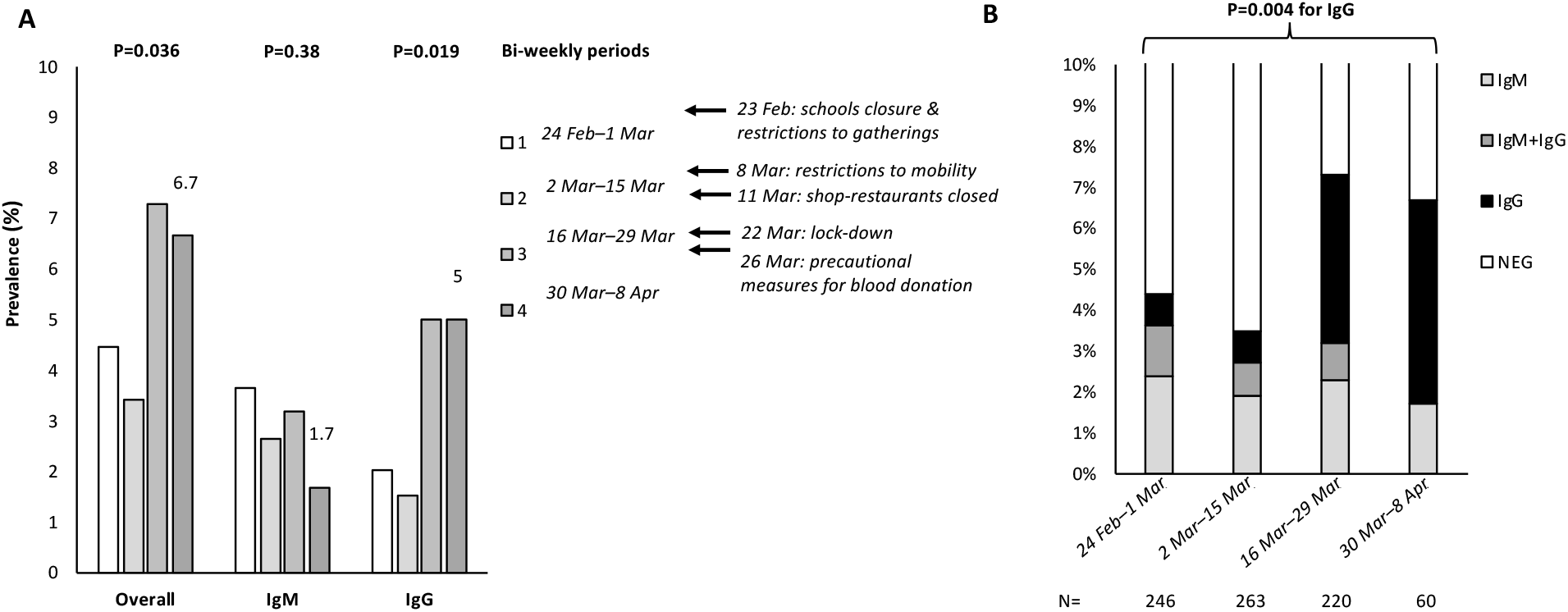
Seroprevalence trends during the COVID-19 Milan outbreak and lock-down. **A)** Overall seropositivity, IgM+ and IgG+ trends in 789 healthy blood donors enrolled in the CoDS study, stratified by the time of evaluation (bi-weekly periods). *P* values were adjusted for age, sex, and BMI. Main political measures to limit the contagion have been highlighted in the timeline. **B)** Frequency and pattern of antibody positivity during the study period (n=789).

The evolution of the combined IgM/IgG+ during the study period is presented in Figure 1B. There was a significant trend for an increase in IgG+ over time (*P*=0.004). At multivariate logistic regression analysis, adjusted for age, sex and BMI, the rate of seroconversion to IgG+ in the study cohort was 2.7±1.3% per week (*P*=0.005). During the last three weeks of the study (last two periods in Figure 1) the prevalence of IgG+ was 5%, 95% c.i. 2.8-8.2%, and the overall prevalence of infection 7.1%, 95% c.i. 4.4-10.8. After adjustment for imperfect accuracy of the test, the estimated prevalence was 5.2%, 95% c.i. 2.4-9.0

At the level of the province of Milan, these estimates would correspond at April 8^th^ to 169,520 undiagnosed cases (3,260,000*0.052), meaning that only one in 15.1 (6.6%: 12,039/(12,039+231,460) as reported by the Italian Ministry of Health) were diagnosed.

The seroprevalence during the 3 months preceding the study is reported in the supplementary results and Figure S2.

By using a Bayesian approach considering a wide range of test performance, the estimated mode of the prevalence during the first two weeks was 1.0%, 95% c.i. 0.1-5.6%, whereas during the last three weeks was 4.5%, 95% c.i. 0.9-9.2%. The probability distribution is shown in Figure S3.

### Clinical features of seropositive individuals

The predictors of a serological pattern suggestive of SARS-CoV-2 previous infection (IgG+) and recent infection (IgM+) are shown in Table 2. IgG+ increased progressively with time (*P*=0.039), and was more frequently detected in younger individuals (*P*=0.043). After adjustment for age, sex, and BMI, IgG+ was not associated with altered laboratory parameters (not shown).

**Table 2.**
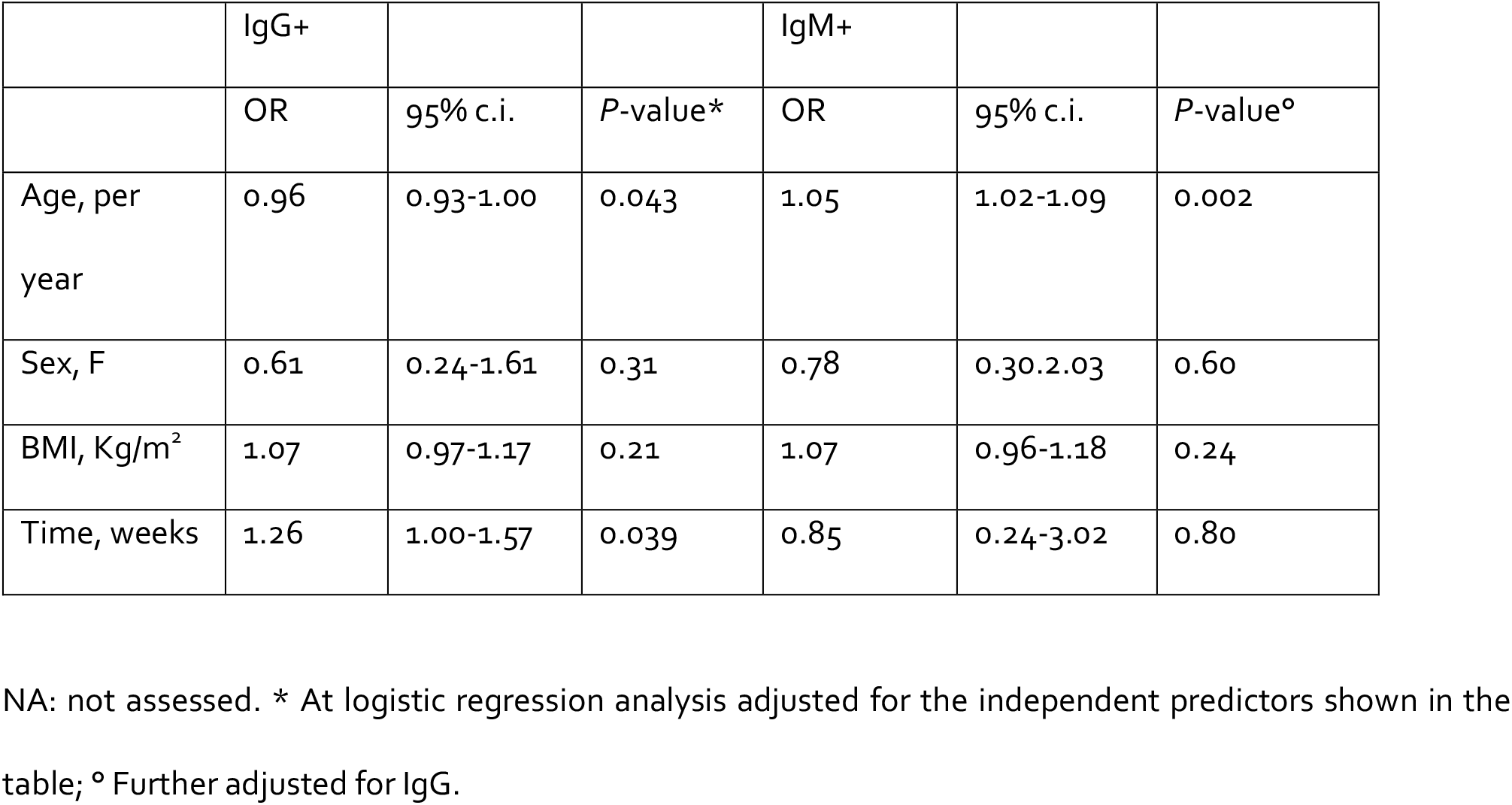
Independent predictors of the presence of anti-SARS-CoV-2 IgG and IgM antibodies in 789 healthy individuals, who donated blood between February 24^th^ and April 8^th^.

|  | IgG+ |  |  | IgM+ |  |  |
| --- | --- | --- | --- | --- | --- | --- |
|  | OR | 95% c.i. | P-value* | OR | 95% c.i. | P-value° |
| Age, per year | 0.96 | 0.93-1.00 | 0.043 | 1.05 | 1.02-1.09 | 0.002 |
| Sex, F | 0.61 | 0.24-1.61 | 0.31 | 0.78 | 0.30-2.03 | 0.60 |
| BMI, Kg/m<sup>2</sup> | 1.07 | 0.97-1.17 | 0.21 | 1.07 | 0.96-1.18 | 0.24 |
| Time, weeks | 1.26 | 1.00-1.57 | 0.039 | 0.85 | 0.24-3.02 | 0.80 |

NA: not assessed. \* At logistic regression analysis adjusted for the independent predictors shown in the table; ° Further adjusted for IgG.

IgM+ increased with older age (p=0.002), but did not change with time. In particular, IgM+ was more frequently observed in donors older than 45 years (18/331, 5.4% vs. 6/457, 1.3%, *P*=0.001). At logistic regression analysis adjusted for age, sex, BMI and IgG+ (Table 3), IgM+ was associated with higher triglycerides (*P*=0.0078), circulating eosinophils (*P*=0.036), and lymphocytes (*P*=0.048).

**Table 3.**
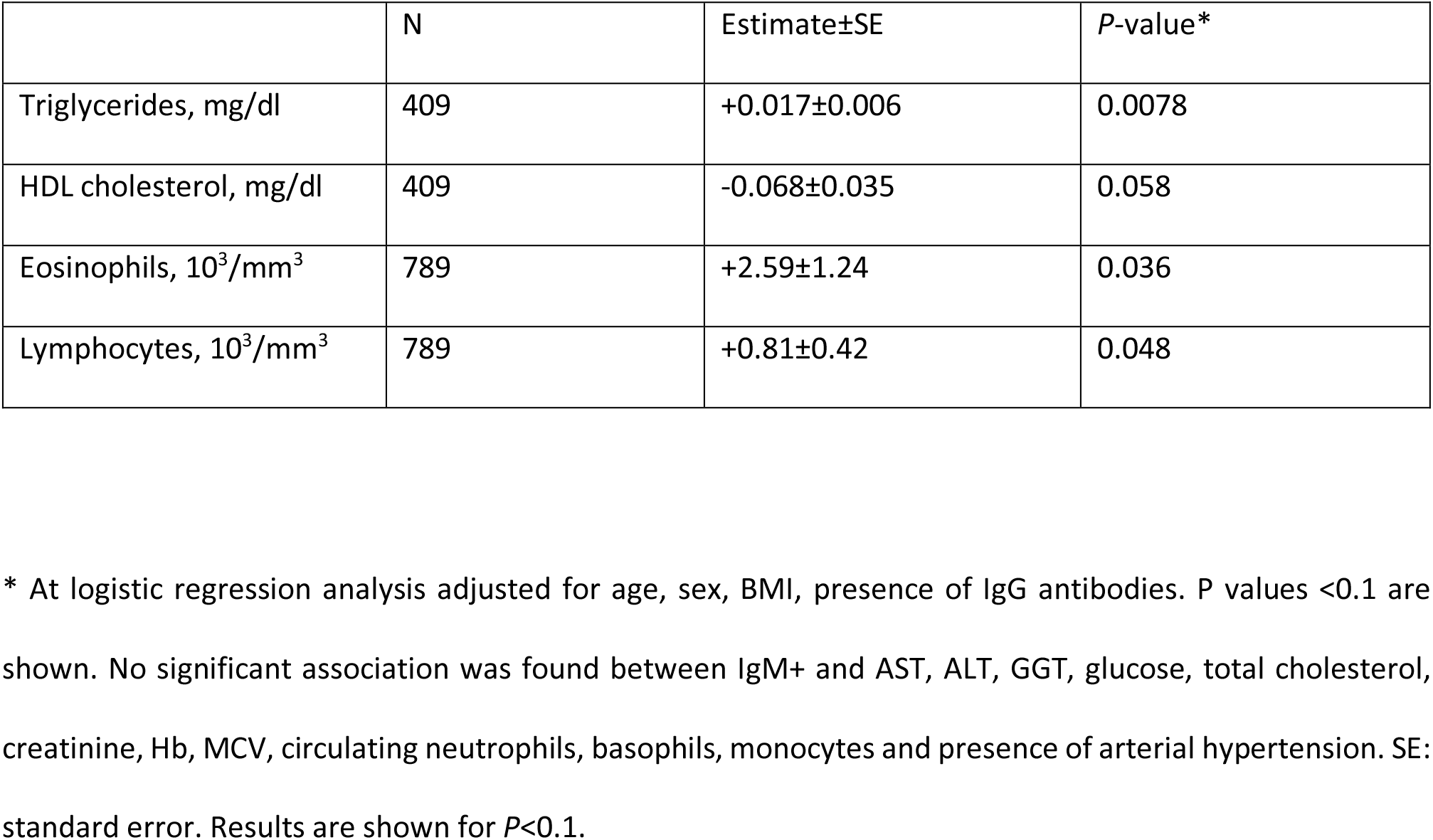
Biochemical and hematological parameters associated with detection of IgM antibodies (n=24) against SARS-CoV-2 among 789 healthy individuals, who donated blood between February 24^th^ and April 8^th^.

|  | N | Estimate±SE | P-value* |
| --- | --- | --- | --- |
| Triglycerides, mg/dl | 409 | +0.017±0.006 | 0.0078 |
| HDL cholesterol, mg/dl | 409 | -0.068±0.035 | 0.058 |
| Eosinophils, 10<sup>3</sup>/mm<sup>3</sup> | 789 | +2.59±1.24 | 0.036 |
| Lymphocytes, 10<sup>3</sup>/mm<sup>3</sup> | 789 | +0.81±0.42 | 0.048 |

\* At logistic regression analysis adjusted for age, sex, BMI, presence of IgG antibodies. P values <0.1 are shown. No significant association was found between IgM+ and AST, ALT, GGT, glucose, total cholesterol, creatinine, Hb, MCV, circulating neutrophils, basophils, monocytes and presence of arterial hypertension. SE: standard error. Results are shown for $P < 0.1$ .

## DISCUSSION

In this study, we examined the SARS-CoV-2 seroprevalence trend in a random sample of healthy blood donors during the Milan COVID-19 outbreak. We exploited a lateral membrane immunoassay that showed an acceptable specificity and concordance with and independent ELISA test.

The first finding was that during the last week of February, considered the start of the outbreak in the Lombardy region around Milan [13], about 4.6% of healthy adults were already positive for IgM (3.7%) and/or IgG (2.0%) antibodies, leading to an estimated true prevalence of SARS-CoV-2 infection of about 2.7% (1.0% by a Bayesian approach). These data indicate that the infection was spreading in the population before the rapid rise in severe COVID-19 cases was observed. During the study period, we observed an adjusted rate of increase in IgG+ of 2.7±1.3% per week. However, seroconversion to IgG+ likely reflected infections acquired before major social distancing measures were enacted.

The divergent impact of age on seroprevalence trends (that is, IgG associated with younger and IgM with older age) is consistent with the possibility that before the restrictions SARS-CoV-2 spread was more diffuse in younger individuals, whereas after closure of schools and universities the spread was mainly supported by work contacts among senior active individuals. These residual infections may account for the lack of a significant decrease in IgM+, despite a non-significant trend, during the observation period. Alternatively, residual IgM+ may also be accounted by false positive results. These data are consistent with a favorable impact of school closure and the lock-down on disease spread [13]. In contrast, we did not observe a significant impact of carrying the A blood group on IgM/G+ [14].

Interestingly, we detected an association between a serological pattern consistent with recent infection (IgM+) and higher triglycerides, eosinophils and lymphocytes. Hypertriglyceridemia has been associated with inflammation in patients with COVID-19, but severe infection is usually associated with a decrease in circulating lymphocytes and eosinophils [15, [16]. It could be speculated that an effective immune reaction against SARS-COV-2 is marked by a distinct pattern of immune and circulating leukocytes response. Of note, eosinophils have recently been implicated in the mucosal response to viral infections in the lung and the immune homeostasis and IgA production in the intestine [17]. However, the interpretation of these findings is limited by the fact that given the diagnostic accuracy of the test and disease prevalence, in more than one third of cases positive tests are likely not specific.

This study has other limitations. Lateral flow immunoassays may have limited accuracy, although they showed adequate performance for epidemiological studies [18]. Although follow-up data are not yet available, the test we used showed was validated in a subset of cases by ELISA. In addition, an overestimation of the sensitivity of the test may have led to a modest underestimation of the true prevalence of SARS-CoV-2 infection, but the adjusted rate of IgG+ increase during the study was less likely affected. Blood donors are generally healthier than the general population, therefore they might have a higher number of social interactions than other groups. Furthermore, they do not include extreme age groups, at different risk of severe COVID-19.

In conclusion, SARS-CoV-2 infection was already circulating in Milan at the start of COVID-19 outbreak. Social distancing may have been more effective in younger individuals, and by April 8^th^ 2020 2.4-9.0% of healthy adults had evidence of seroconversion. Asymptomatic infection may have an impact on laboratory tests.

## Data Availability

The study database is available upon request for collaborative studies.

## SUPPLEMENTARY MATERIAL

INDEX

Page 4: Supplementary results

Page 5: Supplementary figures

## SUPPLEMENTARY RESULTS

### Seroprevalence trends since December 2019

In a subgroup of donors older than 40 years with dysmetabolism, and in donors selected according to similar criteria (older than 40 years and with BMI≥25 Kg/m^2^) are shown in Figure S2A. Between December 2019 and March 2020, the prevalence of infection was 5/184: 2.7%, 95% c.i. 1.2-6.2%. A similar trend for an increase in the prevalence of IgG+ was also observed in donors with dysmetabolism during the study period. The first IgM+ test was detected on December 9^th^ 2019, while the first IgG+ test on December 11^th^. The overall trend of antibody prevalence in all donors stratified by month is shown in Figure S2B.

## SUPPLEMENTARY FIGURES

**Figure S1.**
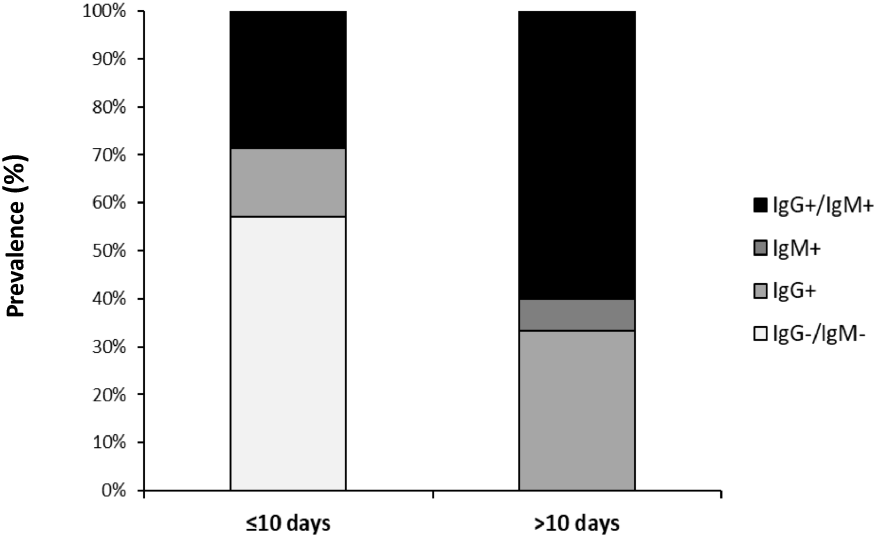
Frequency of positivity for IgG + alone, IgM + alone, IgG and IgM + in patients tested before and after ten days since the onset of symptoms.

**Figure S2.**
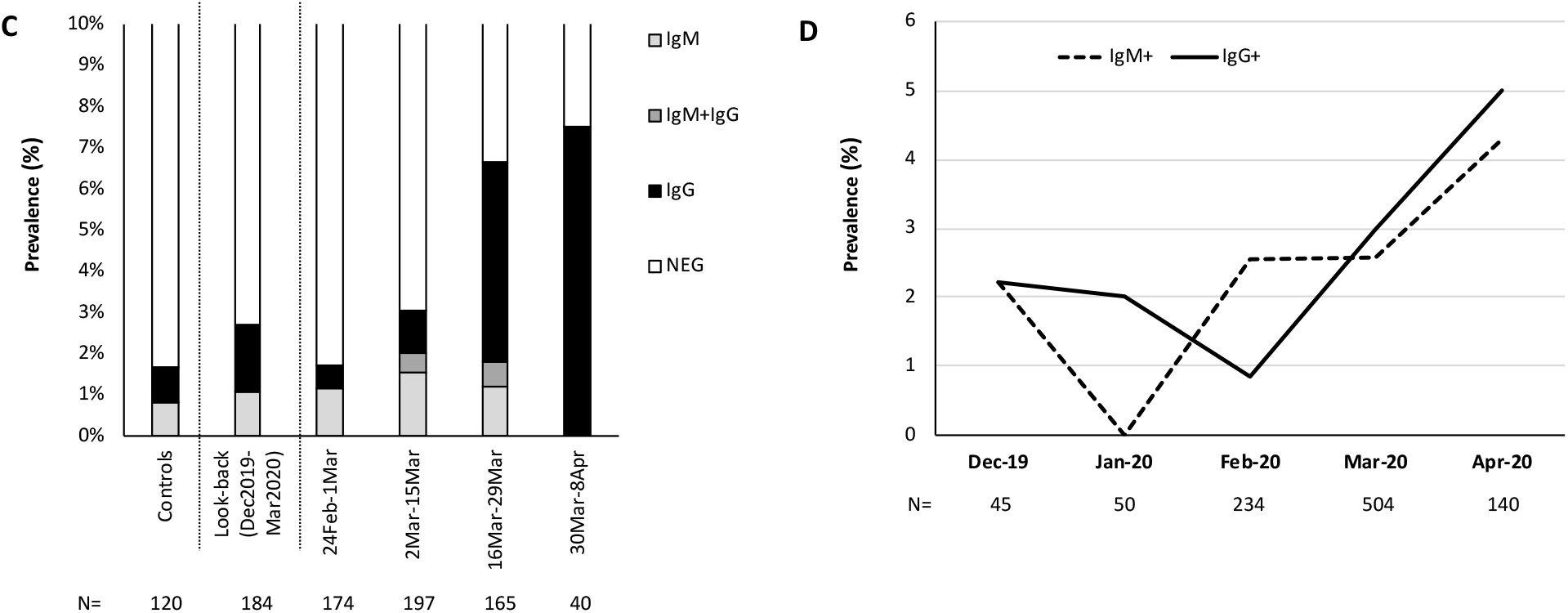
**A**) Frequency and pattern of antibody positivity during the preceding months in 184 individuals older than 40 years with dysmetabolism, and in a comparable subgroup of blood donors during the study period. Historical controls are shown as reference for the specificity of the test. **B**) Trends in IgM+ and IgG+ in 973 donors stratified by month of evaluation.

**Figure S3.**
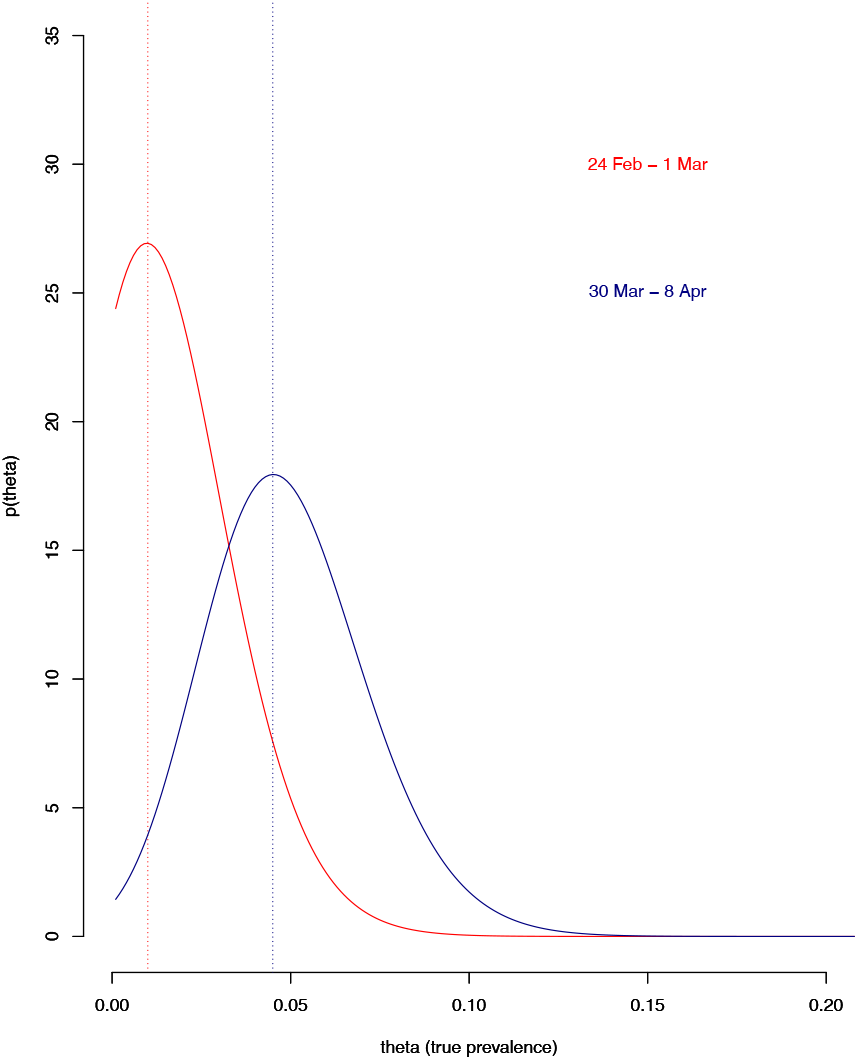
Probability distribution of SARS-CoV-2 prevalence at the beginning (24 Feb – 1 Mar, red curve) and end (30 Mar – 8 Apr, blue curve) of the study period, estimated by a Bayesian approach. Dotted lines represent the mode of the distribution.

